# A genome-wide meta-analysis of palmoplantar pustulosis implicates Th2 responses and cigarette smoking in disease pathogenesis

**DOI:** 10.1101/2024.01.17.24301406

**Authors:** Ariana Hernandez-Cordero, Laurent Thomas, Alice Smail, Zhao Qin Lim, Jake R Saklatvala, Raymond Chung, Charles J Curtis, Patrick Baum, Sudha Visvanathan, A David Burden, Hywel L Cooper, Giles Dunnill, Christopher EM Griffiths, Nick J Levell, Richard Parslew, Nick J Reynolds, Shyamal Wahie, Richard B Warren, Andrew Wright, The APRICOT and PLUM Study Team, Michael Simpson, Kristian Hveem, Jonathan N Barker, Nick Dand, Mari Loset, Catherine H Smith, Francesca Capon

## Abstract

**Background:** Palmoplantar pustulosis (PPP) is an inflammatory skin disorder that mostly affects smokers and manifests with painful pustular eruptions on the palms and soles. While the disease can present with concurrent plaque psoriasis, TNF and IL-17/IL-23 inhibitors show limited efficacy. There is therefore a pressing need to uncover PPP disease drivers and therapeutic targets.

**Objectives:** To identify genetic determinants of PPP and investigate whether cigarette smoking contributes to disease pathogenesis.

**Methods:** We performed a genome-wide association meta-analysis of three North-European cohorts (n=1,456 PPP cases and 402,050 controls). We then used the scGWAS program to investigate the cell-type specificity of the resulting association signals. We undertook genetic correlation analyses to examine the similarities between PPP and other immune-mediated diseases. Finally, we applied Mendelian randomization to analyze the causal relationship between cigarette smoking and PPP.

**Results:** We found that PPP is not associated with the main genetic determinants of plaque psoriasis. Conversely, we identified genome-wide significant associations with the *FCGR3A/FCGR3B* and *CCHCR1* loci. We also observed 13 suggestive (*P*<5X10^-6^) susceptibility regions, including the *IL4/IL13* interval. Accordingly, we demonstrated a significant genetic correlation between PPP and Th2-mediated diseases like atopic dermatitis and ulcerative colitis. We also found that genes mapping to PPP-associated intervals were preferentially expressed in dendritic cells and enriched for T-cell activation pathways. Finally, we undertook a Mendelian randomization analysis, which supported a causal role of cigarette smoking in PPP.

**Conclusions:** The first genome-wide association study of PPP points to a pathogenic role for deregulated Th2 responses and cigarette smoking.

**Clinical implications:** The results of the first PPP GWAS support the therapeutic potential of agents that inhibit Th2 responses and target inflammatory pathways activated by cigarette smoking.

**Capsule:** The genetic analysis of ∼1,400 PPP cases and 400,000 healthy controls points to a causal role of abnormal Th2 responses and cigarette smoking. This supports the therapeutic utility of Th2 inhibition.

## Introduction

Palmar plantar pustulosis (PPP) is a severe inflammatory skin disorder presenting with neutrophil-filled pustules on the palms and soles^1^. The lesions are painful, disabling and stigmatizing, so that PPP has a profound impact on quality of life. This is often compounded by comorbidities such as type 1 diabetes, psoriatic arthritis and Graves’ disease^2^. At the same time, PPP remains very difficult to manage, with treatment options limited by poor efficacy and/or toxicity^3^.

As PPP is traditionally classified as a pustular variant of psoriasis, clinical trials have tested the efficacy of agents used to good effect in plaque psoriasis (TNF, interleukin [IL]-17 and IL-23 blockers) or generalized pustular psoriasis (IL-1 and IL-36 blockers). These therapeutics, however, showed limited efficacy in PPP is limited^4-7^.

These disappointing results reflect a poor understanding of the pathways underlying disease pathogenesis. The analysis of candidate genes and bulk RNA-sequencing profiles has documented a significant up-regulation of Th17 responses in PPP^8-10^. This was confirmed in a recent single-cell study carried out by our group, which also demonstrated an abnormal activation of Th2 pathways^10^. It is, however, unclear whether these alterations are a cause or a secondary manifestation of the disease. PPP disproportionately affects women^11, 12^ and smokers ^11, 12^, with evidence of increased disease severity in current/heavy smokers vs former/never smokers^13-15^. While it has been hypothesized that nicotine alters the expression of IL-36 cytokines^14^ and anti-inflammatory nicotinic acetylcholine receptors^16^, a causal role of cigarette smoking has not been established in PPP.

Genome-wide association studies (GWAS) and Mendelian randomization approaches have unique potential to reveal pathogenic pathways and disease-promoting exposures, especially when applied to large population biobanks^17^. This promise, however, has not been realized in PPP, as the disease is poorly annotated in the UK Biobank (<50 cases reported) and its rarity has hindered the ascertainment of adequately powered datasets.

Here, we have addressed these issues by combining three independent Northern European cohorts, including a total of 1,456 cases and 402,050 controls. By undertaking a genome-wide association meta-analysis in this extended dataset, we identified two genome-wide significant and 13 suggestive susceptibility loci. We then investigated our association results by genetic correlation analysis and Mendelian randomization, implicating Th2 responses and cigarette smoking in the pathogenesis of PPP.

## Methods

### Study cohorts

This work was undertaken according to the principles of the Declaration of Helsinki. Ethical approval was obtained from the Norwegian Data Protection Authority and the Regional Committee for Medical and Health Research Ethics in Central Norway (Reference number 2015/586); London Bridge Research Ethics Committee, UK (ref: 16/LO/2190) and the Coordinating Ethics Committee of the Hospital District of Helsinki and Uusimaa (ref: HUS/990/2017). Written informed consent was obtained from all study participants.

Three patient cohorts, originating from the UK, Norway and Finland, were examined. The UK dataset included 288 affected individuals of European descent, who were recruited in specialist dermatology centers as part of the APRICOT clinical trial^4^ and its sister study PLUM. All were diagnosed by dermatologists, based on clinical examination and consensus criteria^18^. A total of 7,321 unrelated individuals from the English Longitudinal Study of Aging were analyzed as controls.

The Norwegian cases (n=225) were ascertained from the Trøndelag Health Study (HUNT), a population-based study of adult residents from Trøndelag County^19^. Participant IDs were linked to regional- and national health registries, which enabled the identification of PPP cases through the L40.3 (pustulosis palmaris et plantaris) ICD-10 code. HUNT participants who were not affected by PPP or any other form of psoriasis (n=64,050) were analysed as controls.

The Finnish sample was also a population-based cohort. It was derived from the 8^th^ data release of the FinnGen study, a partnership aiming to analyse genome and health data from Finnish biobank participants^20^. Cases (n=969) were ascertained based on the L40.3 ICD-10 code and the remaining FinnGen participants (n=330,975) were analysed as controls. Key demographic and clinical information for the PPP cases in the three cohorts is summarised in Table E1.

### Effect of disease associated SNPs on gene expression

Following genotyping, association testing and meta-analysis (all described in the Supplementary methods), the potential effects of PPP associated SNPs on gene expression were investigated. Summary data-based Mendelian randomization was therefore implemented with SMR v1.3.1^21^. Summary statistics for expression quantitative trait loci (eQTL) identified in non-sun exposed skin were retrieved from the GTEx database (V7 release, https://gtexportal.org/home/datasets) and examined in conjunction with the PPP meta-analysis results. The genome-wide threshold for statistical significance was set at *P*<9.6x10^-6^ (0.05/5,189 probes). To determine whether gene expression and disease phenotype were influenced by a shared variant (pleiotropy) or multiple variants in linkage disequilibrium (LD) with each other (linkage), a HEIDI test (heterogeneity in dependent instruments) was undertaken. Any SNP generating a statistically significant SMR p-value and a HEIDI p-value >0.05 was deemed to have pleiotropic effects on PPP and gene expression.

### scRNA-seq assisted GWAS analysis (scGWAS)

We used scGWAS_r1^22^ to determine if the PPP-associated genes identified in the GWAS are concordantly activated in a particular cell type. We first calculated gene-based p-values by processing the meta-analysis summary statistics with MAGMA^23^ (Multi-marker Analysis of GenoMic Annotation). The programme aggregates the SNP data available for each gene and implements a multiple regression analysis to test their joint association with the phenotype of interest. Next, we obtained two reference scRNA-seq datasets: i) the peripheral blood mononuclear cells (PBMC) profiles provided by Jia et al (n=1 donor)^22^ and ii) a subset of the healthy PBMC profiles generated by McCluskey et al (n=3 donors)^10^, which we re-analysed as described in the Supplementary Methods. We applied the Box-Cox transformation to the original distribution of -log10 (p) from MAGMA and the distribution of log (CPM+1) from the scRNA-seq datasets. Finally, we processed the two normalised datasets with scGWAS.

### Genetic covariance and pathway enrichment analysis

We used GNOVA (GeNetic cOVariance Analyzer)^24^ to investigate pairwise genetic correlations between PPP and traits of interest (plaque psoriasis and four diseases where the involvement of Th2 pathways is well recognised: allergic rhinitis, asthma, atopic dermatitis (AD), and ulcerative colitis (UC)). We retrieved summary statistics from studies shared by the UK biobank (allergic rhinitis; dataset: ukb-b-16499) or published in the literature (all other diseases^25-28^). We removed low-frequency (minor allele frequency <5%) and palindromic SNPs. We then calculated genetic correlations, using the European 1000 Genomes Phase 3^29^ (v5) dataset as a reference panel for LD estimation. We set the threshold for statistical significance at *P*<0.01 (0.05/5 traits). For pathway enrichment analyses, we processed GWAS summary statistics with MAGMA to obtain gene-based p-values. We ranked the output gene list by p-value and analysed it with GSEA^30^, using Gene Ontology Biological Process terms as a reference dataset. Network analysis and visualization was then undertaken with Cytoscape^31^ 3.10.1.

### Mendelian randomization

To investigate the causality of cigarette smoking in PPP, Mendelian randomization was implemented with TwoSampleMR v0.5.7^32^. The genetic instrument recapitulating the effects of the exposure was derived from a GWAS for smoking initiation undertaken in 3,383,199 individuals^33^. Clumping of associated variants was performed to only keep one SNP per LD block (LD window=500kb, r^2^=0.01). If a lead SNP from the exposure GWAS was not present in the outcome dataset (our PPP meta-analysis), a proxy with r^2^>0.5 was identified with LDproxy v5.6.3^34^, using the European 1000 Genomes Phase 3 dataset as a reference. Finally, palindromic SNPs were removed and the power of the resulting genetic instrument (n=240 markers) was validated by calculating the F-statistic. As F was 38.1, we were able to exclude weak instrument bias (observed when F<10) and implement Mendelian randomization with the inverse variance weighted and weighted median methods. To validate the robustness of our findings, additional tests were performed to examine heterogeneity (inverse variance weighted method), horizontal pleiotropy (Egger intercept) and direction of effect (r^2^ exposure vs r^2^ outcome). The effects of individual SNPs were also examined to investigate sensitivity.

## Results

### Genome-wide meta-analysis

To identify genetic determinants of PPP we undertook a genome-wide association scan of 288 UK cases and 7,321 unaffected controls. We then used METAL^35^ to meta-analyze our results with those obtained in two population-based cohorts analyzed by the HUNT study (225 cases and 64,050 controls) and the FinnGen consortium (969 cases and 330,975 controls). The total dataset included 403,506 individuals (1,456 cases and 402,050 controls), genotyped for ∼9.2M SNPs (Table E1). The distribution of P-values did not deviate from that expected in the absence of population stratification (λ=1.05) (Figure E1).

The meta-analysis yielded two genome-wide significant loci, spanning *CCHCR1/POU5F1* (*P*=2.9x10^-11^) and *FCGR3A/3B* (1.6 x10^-8^) (Figure 1; Figure E2). Of note, *CCHCR1* maps in close proximity of *HLA-C*0602*, the major genetic determinant of plaque psoriasis^36^. To exclude the possibility that the association we had observed was driven by co-morbid plaque psoriasis (reported in 7-28% of our PPP cases) we examined LD conservation around the *CCHR1* locus. By interrogating population data generated in Finnish and British individuals, we found that the lead SNP in this region was not in LD with rs4406273 (r^2^<0.15) (Figure E3), a well-known *HLA-C*0602* proxy^37^. Accordingly, rs4406273 was not associated with PPP in the FinnGen or UK dataset (*P*>0.05 for both; no data was available for HUNT where the SNP had not been typed or imputed). Thus, the *CCHCR1* association is independent of *HLA-C*0602*, which is consistent with LD patterns previously described in other datasets^38^.

**Figure 1:**
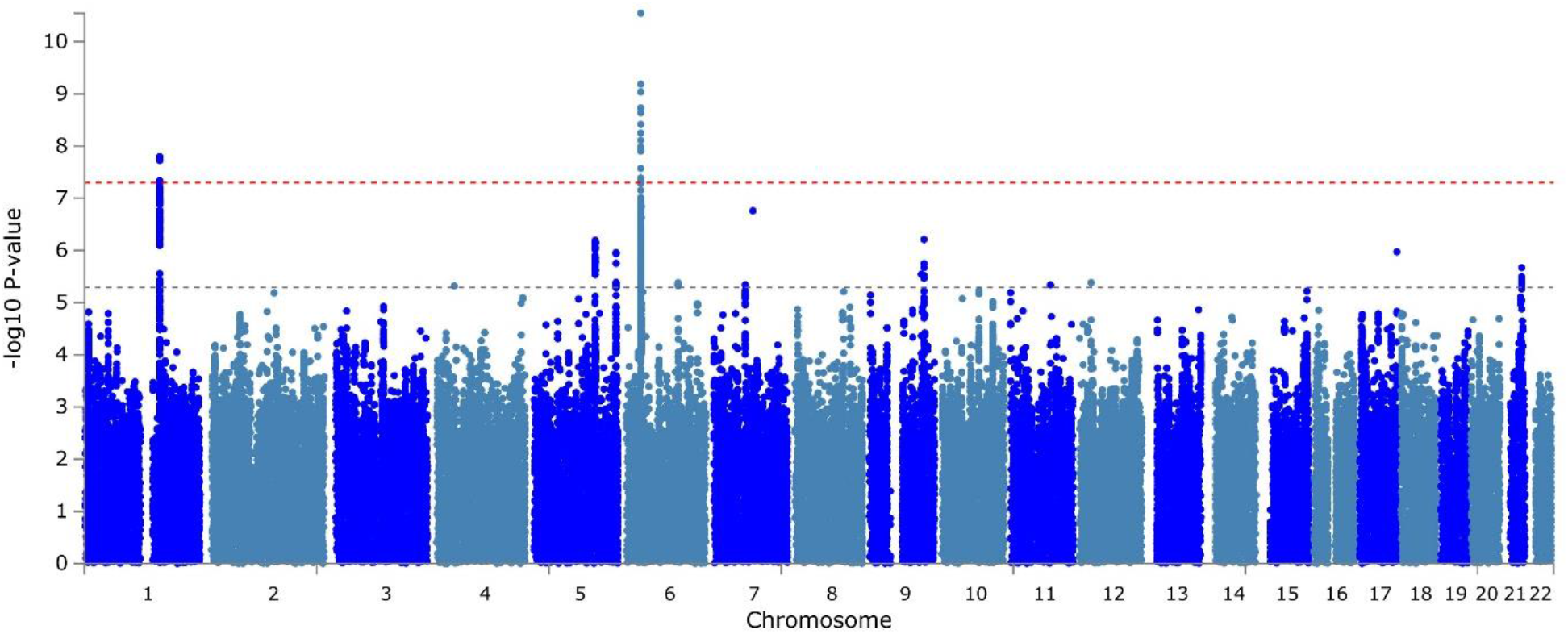
Manhattan plot showing the association signals detected in the GWAS meta-analysis.

We next undertook conditional analysis at the *CCHCR1/POU5F1* and *FCGR3A/FCGR3B* loci. We did not detect any secondary association signals, although this may be due to limited statistical power.

Beyond *CCHCR1/POU5F1* and *FCGR3A/FCGR3B*, a further 13 regions showed suggestive evidence for association, with P-values <5x10^-6^ observed in proximity of immune genes such as *IL4, HLA-DRA* and *TNFSF15* (Table 1, Table E2).

**Table 1:** Summary of genome-wide significant and suggestive association signals.

| rsID | Position<sup>1</sup> | EAF/NEAF | EAF-cases | EAF-controls | P value | OR | 95% CI | Direction | Nearest gene<sup>2</sup> |
| --- | --- | --- | --- | --- | --- | --- | --- | --- | --- |
| rs61802325 | 1:161,588,097 | A/G | 0.6402 | 0.6076 | $1.60 \times 10^{-08}$ | 1.28 | 1.24 - 1.31 | +++ | *FCGR3B* |
| rs887467 | 6:31,141,664 | C/G | 0.4301 | 0.4788 | $2.87 \times 10^{-11}$ | 0.75 | 0.72 - 0.79 | --- | *POU5F1/CCHCR1* |
| rs73236841 | 4:37,911,079 | A/C | 0.7731 | 0.7905 | $4.79 \times 10^{-06}$ | 0.78 | 0.74 - 0.83 | --- | *TBC1D1* |
| rs3798130 | 5:132,042,146 | T/C | 0.3170 | 0.3067 | $6.41 \times 10^{-07}$ | 1.27 | 1.23 - 1.31 | +++ | *KIF3A/IL4* |
| rs4075959 | 5:176,784,612 | A/G | 0.3371 | 0.3001 | $1.10 \times 10^{-06}$ | 1.26 | 1.22 - 1.30 | +++ | *RGS14* |
| rs9487605 | 6:111,582,885 | A/G | 0.3970 | 0.3678 | $4.15 \times 10^{-06}$ | 1.23 | 1.19 - 1.27 | +++ | *MFSD4B* |
| rs2097442 | 6:32,422,191 | A/G | 0.3238 | 0.2787 | $1.43 \times 10^{-07}$ | 1.29 | 1.24 - 1.33 | +++ | *HLA-DRA* |
| rs10950151 | 7:68,306,574 | T/C | 0.0709 | 0.0566 | $4.54 \times 10^{-06}$ | 1.54 | 1.46 - 1.62 | +++ | - |
| rs1990107 | 7:84,724,076 | T/C | 0.0407 | 0.0257 | $1.75 \times 10^{-07}$ | 2.00 | 1.88 - 2.11 | +++ | *SEMA3D* |
| rs11793564 | 9:111,534,315 | A/G | 0.7523 | 0.7099 | $2.87 \times 10^{-06}$ | 1.25 | 1.21 - 1.29 | +++ | *ACTL7B* |
| rs4246905 | 9:117,553,249 | T/C | 0.1913 | 0.2134 | $6.16 \times 10^{-07}$ | 0.78 | 0.73 - 0.82 | --- | *TNFSF15* |
| rs9666271 | 11:85,879,769 | T/C | 0.9738 | 0.9624 | $4.58 \times 10^{-06}$ | 1.64 | 1.54 - 1.73 | +++ | - |
| rs860876 | 12:25,158,319 | T/G | 0.2038 | 0.1720 | $4.14 \times 10^{-06}$ | 1.30 | 1.25 - 1.35 | +++ | *IRAG2* |
| rs112872175 | 17:80,589,968 | G/GATAA | 0.2094 | 0.1843 | $1.06 \times 10^{-06}$ | 1.31 | 1.26 - 1.36 | +++ | *FOXK2/WDR45B* |
| rs4817988 | 21:40,468,838 | A/G | 0.2184 | 0.2541 | $2.15 \times 10^{-06}$ | 0.79 | 0.75 - 0.84 | --- | *PSMG1* |

CI, confidence intervals; EA, effect allele; EAF, effect allele frequency NEAF, non-effect allele frequency; OR, odds ratio. <sup>1</sup>Coordinates refer to GRCh37.
- and + refer to protective and risk-bearing effects in individual cohorts. <sup>2</sup>Nearest gene(s) identified by LocusZoom.

### Analysis of genes and cell types underlying association signals

While the lead SNPs for the *CCHCR1/POU5F1* and *FCGR3A/FCGR3B* loci were in LD with coding variants (rs130068 and rs76714703, respectively; r^2^>0.75 for both) neither of these missense changes was classified as damaging by pathogenicity predictors (CADD^39^ and MutationTaster^40^). We therefore examined the overlap between the two association signals and skin expression quantitative trait loci (eQTL) identified by the GTEX consortium^41^. Using the SMR program, we found evidence of pleiotropy for SNP rs74538320 in the chromosome 1 locus, indicating that the variant affects PPP risk and *FCGR3B* expression (pSMR=5.3x10^-6^; pHEIDI>0.05). Conversely, the analysis of the *CCHCR1/POU5F1* locus identified a disease-associated SNP (rs1265079) that is unlikely to directly influence gene expression, but may be in LD with a variant regulating *CCHR1* levels (pSMR=4.4x10^-6^; pHEIDI=4.5x10^-5^).

We ran a parallel analysis of GTEX blood eQTL, but neither of the above SNPs generated a significant SMR p-value. Of note, we previously showed that circulating immune cells are likely to play a role in the immune pathogenesis of PPP^10^. To further investigate our GWAS results in the light of these observations and determine whether the effects of susceptibility alleles were mediated by specific immune populations, we undertook scRNA-seq assisted GWAS analysis^22^ (scGWAS). scGWAS derives biological networks that are simultaneously enriched with disease associated genes and genes that are transcriptionally active in a specific cell type.

Here, we observed that networks centered around Transmembrane Immune Signaling Adaptor *TYROBP* and *FOXO1* were enriched for genes that are associated with PPP and expressed in dendritic cells (Figure 2).

**Figure 2:**
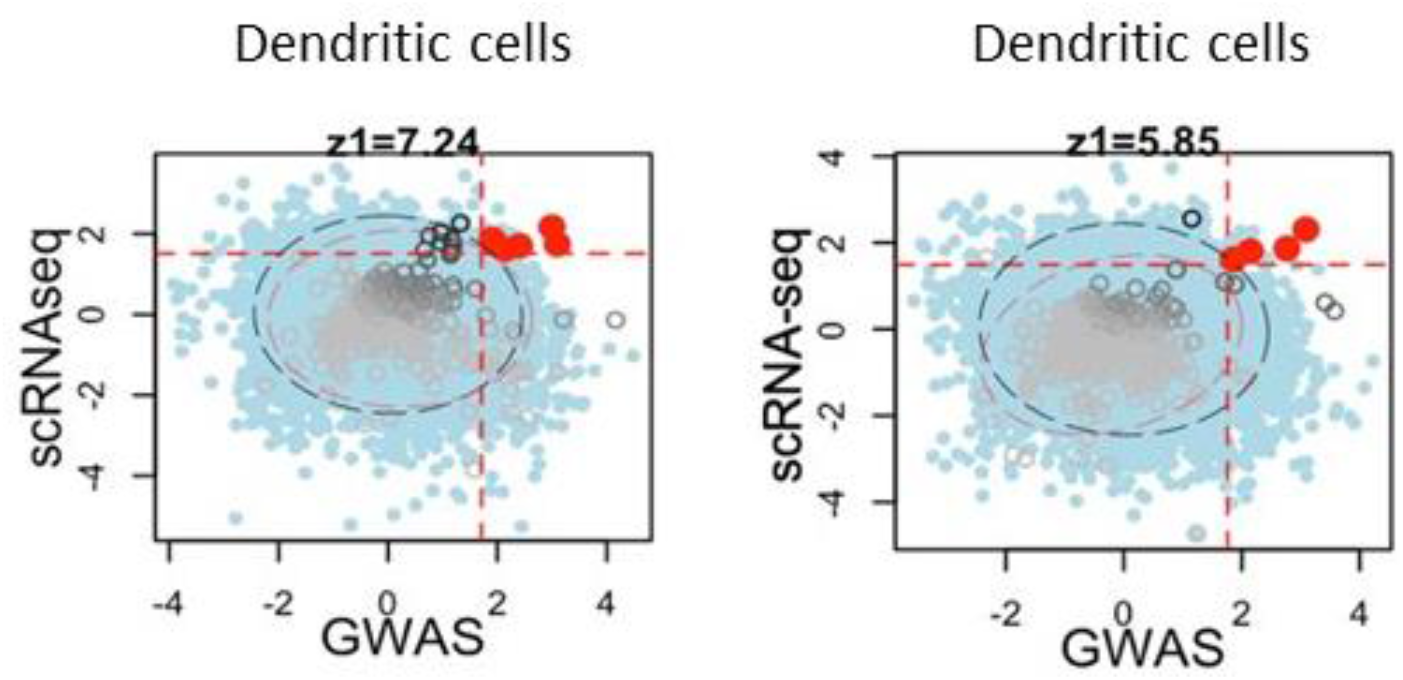
PPP-associated genes are enriched in cell networks (gene modules) that are preferentially expressed in denritic cells. The plots show the results obtained using two scRNA-seq reference dasets for PBMC gene expression. These were retrieved from the scGWAS package (left) and the literature (right, see Methods). The plots show the GWAS (x-axis) and scRNA-seq (y-axis) enrichment scores for each module (dots). Real modules are shown in grey with their confidence intervals indicated by a red circle. Virtual modules, which show the null distribution of module scores, are plotted in blue with confidence intervals in black. The vertical and horizontal dashed lines indicate the enrichment significance threshold (z=1.96) and significant real modules are highlighted in red. The z1 value illustrates the signficance of the red module enrichment.

Taken together, these post-GWAS analyses indicate that the PPP associated alleles affect immune pathways in skin and circulating dendritic cells.

### Genetic correlation between PPP and Th2-mediated diseases

To further investigate the pathophysiological relevance of our findings, we used GNOVA to explore genetic correlations between PPP and other immune traits of interest. Given that PPP can present with concurrent plaque psoriasis^1,13^, we first investigated this condition, examining publicly available GWAS summary statistics^28^. However, we did not find any evidence of correlation between PPP and psoriasis. As we and others have reported Th2 activation in PPP skin^8,10^, we next examined four Th2 mediated disorders (allergy, asthma, AD and UC). We found that PPP shows a positive correlation with AD (r=0.49; *P*=6.1x10^-9^) and an inverse one with UC (r=-0.20; *P*=1.5x10^-4^).

To validate these observations, we undertook pathway enrichment analyses on the GWAS summary statistics generated in PPP, AD, UC and psoriasis. Using GSEA, we identified the 50 most significantly enriched Gene Ontology Biological Processes for each disease (False Discovery Rate <0.05). As expected for the analysis of four immune-mediated disorders, we observed a substantial overlap between the enriched pathways. Importantly, however, the largest number of shared pathways was observed between PPP and UC (Jaccard similarity index=0.33), and PPP and AD (Jaccard similarity index=0.25). A closer inspection of the results revealed that most of the overlapping Gene Ontology processes relate to T-cell differentiation and activation. Conversely, the overlap between PPP and psoriasis was more limited (Jaccard similarity index=0.22) and mostly underpinned by pathways involved in antigen processing/presentation and T-cell cytotoxicity (Figure 3). Taken together, these observations support a shared genetic basis between PPP and Th2 mediated diseases.

**Figure 3:**
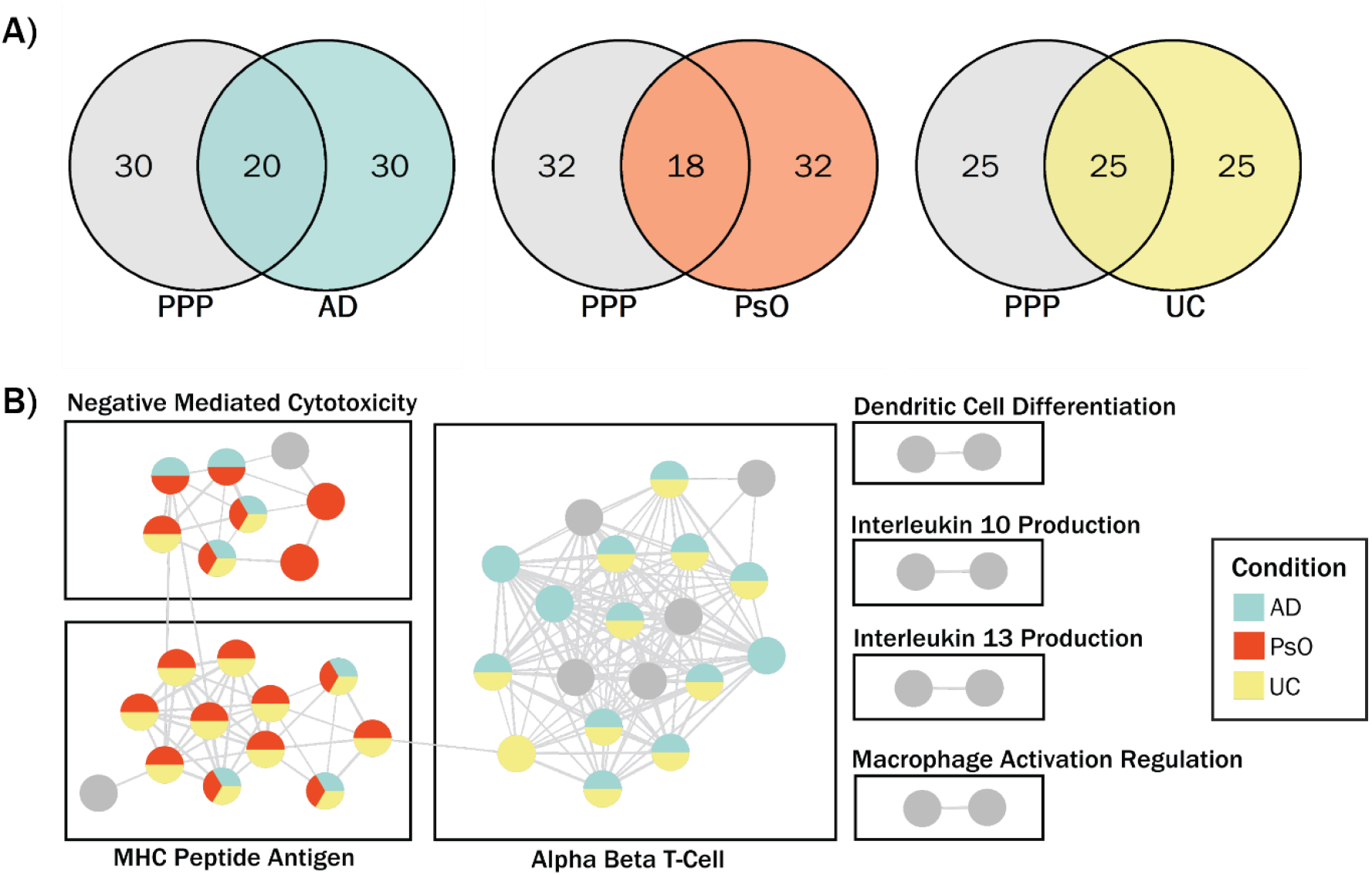
Overlap between GO Biological Process pathways identified from PPP, psoriasis (PsO), AD and UC GWAS summary statistics. **A)** Venn diagrams illustrating the overlap between the 50 most significantly enriched GO Biological Process pathways identified by GSEA in each condition. **B)** Cytoscape network analysis of the 50 most significantly enriched pathways in PPP. Pathways were clustered using the MCL clusterMaker alogrithm and annotated using AutoAnnotate. Each GO pathway is coloured to indicate overlap with AD, PsO and UC; pathways identified in PPP but showing no overlap with AD, PsO or UC are shown in grey.

### Mendelian randomization indicates a causal role of cigarette smoking in PPP

Given the high prevalence of smokers among PPP patients^1,11,13^, we next used Mendelian Randomization to determine whether cigarette smoking may play a causal role in the disease. We first derived a genetic instrument (n=240 independent SNPs associated with smoking initiation^33^) recapitulating the exposure to smoking. We then implemented Mendelian randomization using an inverse variance weighted method. This showed a causal influence of the exposure (smoking) on the outcome (PPP) (*P*=2.9x10^-4^), which was also confirmed with the weighted median method (*P*=1.79x10^-3^).

We tested the robustness of our findings with sensitivity tests measuring heterogeneity between variant causal estimates, horizontal pleiotropy (the phenomenon whereby a SNP may affects the outcome through pathways unrelated to the exposure) and reverse causation. We found no evidence for heterogeneity between variant effects (*P*>0.05), with a leave-one-out test also demonstrating that no SNP had a disproportionate impact on the association between exposure and outcome (Figure E4). We also excluded the presence of horizontal pleiotropy (*P*>0.05) and reverse causation (correct causal direction P-value< 0.001). These results demonstrate that our genetic instrument did not violate key mendelian randomization assumptions, confirming a causal influence of cigarette smoking in PPP.

## Discussion

The aim of this study was to identify novel genetic determinants of PPP through the analysis of an extended dataset. We therefore ascertained three independent case resources and obtained a patient sample of unprecedented size (n=1,456), which enabled us to undertake the first GWAS of PPP.

Our analysis identified two genome-wide significant association signals. The first maps to the *FCGR* cluster on chromosome 1q23, which spans five genes encoding immunoglobulin gamma receptors. Co-localization with skin eQTLs suggested that the association is driven by SNPs downregulating the expression of *FCGR3B*. Interestingly, variation in *FCGR3B* copy number has been associated with susceptibility to systemic lupus erythematosus and rheumatoid arthritis^42,43^. Disease risk is specifically driven by reduced *FCGR3B* copy number, which is thought to impair immunoglobulin binding and clearance of immune complexes^44^. While it is tempting to speculate that a similar mechanism may be at play in PPP, an in-depth dissection of the *FCGR* locus will be required to disentangle the effects of copy number variants and gene eQTLs. The role of the various cell types expressing *FCGR3B* (including T cells, natural killer cells, macrophages and granulocytes) will also require further investigation. Nonetheless, the association between PPP and *FCGR* genes points to an involvement of antibody-mediated pathways, suggesting an autoimmune component for the pathogenesis of PPP.

The second association signal maps to chromosome 6p21. While the lead SNP was located within a *POU5F1* intron, our analysis suggested that it may tag an eQTL for the neighbouring *CCHCR1* gene. The latter encodes a coiled coil protein which has been implicated in microtubule assembly, centrosome maturation, mRNA metabolism and cell proliferation^45^. While a *CCHCR1* coding haplotype has been repeatedly associated with psoriasis susceptibility^46^, the relevant SNPs are not in LD (r^2^<0.5) with the PPP risk allele identified here. This is in keeping with the lack of association observed for the *HLA-C*0602* proxy rs4406273. Of note, we also obtained negative findings (*P*>0.05) for the genomic region encoding *IL36RN*, the main genetic determinant of generalised pustular psoriasis^47^. Thus, PPP is genetically distinct from co-morbid and phenotypically related forms of skin inflammation.

The inspection of suggestive loci revealed that SNPs spanning the *IL4/IL3* gene region, which encodes key Th2 cytokines, are associated with PPP (*P*=6.4x10^-7^ for the lead SNP) and AD^27^. In this context, our GNOVA analysis identified a significant correlation between PPP and AD, suggesting the existence of shared genetic risk. Gene set enrichment analysis also showed that the genes that are associated with both conditions preferentially map to T-cell differentiation and T cell activation pathways. This is in keeping with the notion of a shared T-cell mediated aetiology. The latter is also consistent with the enrichment of PPP associated genes within expression modules centred around *TYROBP* and *FOXO1*. Indeed, both genes are required for T cell activation by dendritic cells^48,49^. Thus, our findings point to a causal role of abnormal Th2 responses in PPP. This observation has significant implications, given that Th2 pathways can be targeted by emerging therapeutics, such as IL-4/IL-13 blockers and JAK1 inhibitors^50^. Interestingly, case reports published in recent months suggest that both class of drugs could have therapeutic efficacy in PPP^51,52^. To complement our genetic findings, we used Mendelian randomization to show that cigarette smoking has a causal influence on PPP. Interestingly, cigarette smoking is thought to be protective in UC^53^, a condition that showed a negative correlation with PPP in the GNOVA analysis. This suggests that the shared genetic determinants between PPP and UC regulate inflammatory pathways that can be influenced by cigarette smoking.

Our Mendelian randomization results are in keeping with reports that disease severity correlates with cigarette pack years^14^. In this context, the further observation that PPP severity is worse in current versus former smokers^13^, suggests that targeting inflammatory pathways influenced by smoking may be an attractive therapeutic strategy.

While ours is by some margin, the largest genetic study of PPP, it is still limited by the relatively small size of the patient sample. The different approaches to case ascertainment (recruitment through Dermatology specialist centres for the UK dataset and analysis of ICD-10 codes for the HUNT and Finngen studies) may have also generated heterogeneity within the case sample, affecting statistical power. Thus, the establishment of larger collaborative consortia will be required to identify further genetic determinants of PPP and validate the involvement of pathways that can be targeted by existing therapeutics.

## Data Availability

The FinnGen data are publicly available. The UK data will be submitted to the GWAS catalogue.

## Abbreviations

AD: atopic dermatitis;
eQTL: expression quantitative trait locus;
GWAS: genome-wide association study;
LD: linkage disequilibrium;
PBMC: peripheral blood mononuclear cells;
PPP: palmoplantar pustulosis;
UC: ulcerative colitis.

## Acknowledgements

We want to acknowledge the participants and investigators of the FinnGen study. In addition to A. David Burden, Hywell L. Cooper, Giles Dunnil, Christopher E. M. Griffiths, Nick J. Levell, Richard Parslew, Nick J Reynolds, Shyamal Wahie, Richard B. Warren, Andrew Wright, Jonathan N. Barker, Catherine H. Smith, and Francesca Capon, who are authors, the following APRICOT and PLUM study team members facilitated patient recruitment and data processing for the APRICOT clinical trial and the PLUM study: Thamir Abraham (Peterborough City Hospital, Peterborough), Muhmad Ali (Worthing Hospital, Worthing, Suzannah August (Poole Hospital NHS Foundation Trust, Poole), David Baudry (Guy’s Hospital, London), Gabrielle Becher (West Glasgow Ambulatory Care Hospital, Glasgow), Anthony Bewley (Whipps Cross Hospital, Leytonstone), Victoria Brown (St Albans City Hospital, St Albans), Victoria Cornelius (Imperial College London, London), Sharizan Ghaffar (Ninewells Hospital and Medical School, Dundee), John Ingram (University Hospital of Wales, Cardiff), Svetlana Kavakleiva (Royal Lancaster Infirmary, Lancaster), Susan Kelly (The Royal Shrewsbury Hospital, Shrewsbury), Mohsen Khorshid (Basildon Hospital, Essex), Helen Lachmann (Royal Free Hospital, London), Effie Ladoyanni (Russells Hall Hospital, Dudley), Helen McAteer (Psoriasis Association, Northampton), John McKenna (Leicester Royal Infirmary, Leicester), Freya Meynell (Guy’s Hospital, London), Prakash Patel (Guy’s Hospital, London), Andrew Pink (Guy’s Hospital, London), Kingsley Powell (Guy’s Hospital, London), Angela Pushparajah (Guy’s Hospital, London), Catriona Sinclair (Broomfield Hospital, Essex) and Rachel Wachsmuth (Royal Devon and Exeter NHS Foundation Trust, Exeter).

