## Supplementary Material for "A genome-wide meta-analysis of palmoplantar pustulosis implicates Th2 responses and cigarette smoking in disease pathogenesis"

**Supplementary Methods**

*Genotyping and data quality control (QC)*

The UK PPP samples were typed on an Infinium Global Screening Array 24 v3.0 (Illumina) and genotypes were called with GenomeStudio v2.0.50 (Illumina). For sample QC, individuals were excluded from further analysis if they had <99% genotype calls, their genotypes did not match the reported sex, they had a high kinship coefficient (>0.088) with another study participant or a heterozygosity ratio deviating by more than three standard deviations from the mean. For genotype QC, SNPs with Hardy-Weinberg equilibrium p-values <10^-10^, call rates <99% or minor allele frequency (MAF) <1% were excluded.

Control UK genotypes from the ELSA cohort (previously generated on lllumina HumanOmni2.5-4v1 and HumanOmni2.5-8v1.3 arrays) were retrieved from the European Phenome-genome archive (https://ega-archive.org/) and subjected to the QC steps described above. Cases and controls were then merged, and principal component (PC) analysis was implemented to exclude individuals of non-European descent.

Norwegian participants from the HUNT2 (1995-1997) and HUNT3 (2006-2008) cohorts were genotyped using HumanCoreExome12 v1.0, HumanCoreExome12 v1.1 or UM HUNT Biobank v1.0 Illumina arrays. Genotypes were called with GenomeStudio v.2011.1 (Illumina). Samples with <99% genotype calls, large copy number variants or >2.5% contamination (as estimated with BAF Regress^1^) were excluded. Individuals of non-European ancestry were also removed, alongside those with X chromosome genotypes that did not match the recorded sex. Rare genetic variants (MAF<1%) and those out of Hardy-Weinberg equilibrium (p-value <0.0001) were excluded.

The FinnGen samples (r8 release) had been typed on Illumina or Affymetrix custom arrays, with calls derived using GenCall/zCall (Illumina) for the former and AxiomGT1 (Applied Biosystems) for the latter. For sample QC, individuals were excluded from further analysis if their genotype did not match the recorded sex, genotype missingness was>5%, heterozygosity deviated from the mean by more than four standard deviations, or non-Finnish ancestry was detected. For genotype QC, variants with call rates <98%, Hardy-Weinberg equilibrium p-values <10^-6^ or minor allele count <3 were excluded.

*Imputation and genome-wide association analysis (GWAS)*

For the UK dataset, the Michigan Imputation Server (https://imputationserver.sph.umich.edu/index.html#!) was used to phase SNPs with Eagle 2.3^2^ and impute genotypes with Minimac4 v4.1.2^3^. The European 1000 Genomes Phase 3 (v5) dataset was selected as a reference panel. The GWAS was undertaken with PLINK 1.94.1^4^. A logistic regression model was applied whereby sex and the first 10 ancestry PCs were analysed as covariates and genotype probabilities (dosages) were used for imputed variants.

For the HUNT study, imputation was performed by phasing SNPs with EAGLE v 2.3 and either 1) imputing genotypes using Minimac3 v2.0.) and using a reference panel constructed from Haplotype Reference Consortium genotypes (release v1.1) and whole-genome sequences of 2202 HUNT participants or 2) imputing genotypes using Minimac4 v1.0.2 and using the European 1000 Genomes Phase 3 (version 5) dataset as a reference panel. The GWAS was run by fitting a logistic mixed model with SAIGE v0.35.8.3^5^, using sex, birth year, genotyping batch and four ancestry PCs as covariates. Genotype dosages were used for imputed variants. All analyses were performed in digital labs at HUNT Cloud, NTNU - Norwegian University of Science and Technology, Trondheim, Norway.

For the FinnGen dataset, imputation was carried using Beagle 4.1^6^ and a population-specific reference panel (Sequencing Initiative Suomi v3, based on whole-genome sequences of 3,775 Finnish individuals), as described elsewhere^7^. The GWAS was implemented with SAIGE v0.35.8.8, using sex, age, genotyping batch and 10 PCs as covariates. Summary statistics were made publicly available and were retrieved for this study through the FinnGen portal (www.finngen.fi/en/access_results).

*Meta-analysis, conditional analysis and linkage disequilibrium (LD) estimates*

A total of 9,168,215 SNPs present in all three datasets and aligned to the same reference allele, were analyzed in 1,456 cases and 402,050 controls. METAL^8^ v2011.03.25 was used to perform a fixed-effect meta-analysis weighted for effective sample size. The associations yielding p-values <5.0×10^-8^ were considered genome-wide significant, whereas those generating p-values <5x10^-6^ were deemed suggestive. The quantile-quantile plot, Manhattan plot and regional association plots were generated with FUMA v1.5.4^9^.

Conditional analysis was performed with GCTA-COJO^10^ v1.94.1, using the UK cohort as a reference sample for LD estimation. P-values <5×10^−8^ were considered genome-wide significant.

LD conservation across the 130 kb region spanning *HLA-C, CCHCR1* and *PSORS1C3* was assessed using the 1000 Genomes Phase 3 genotypes for British and Finnish participants (GBR and FIN datasets). r^2^ correlation coefficients were calculated for all pairs of common SNPs and the results were visualised using Haploview v4.2.0^11^.

*PBMC scRNA-seq re-analysis*

scRNA-seq raw counts were retrieved for three of the healthy PBMC samples analysed by McCluskey et al^12^ (subseries GSE185857; sample ids: HC11, HC13, HC14) and imported in Seurat v4.3.0^13^. Cells with <300 or >5000 gene counts were excluded, alongside cells with >20% mitochondrial gene reads. The three filtered datasets were merged and integrated in a single Seurat object. The data was normalised and then scaled. Following PC analysis, k-nearest neighbourhoods were defined and unsupervised clustering was performed with a resolution of 0.4. The cell clusters were visualised using uniform manifold approximation projection (UMAP) and annotated based on the expression of canonical marker genes.

**Table E1**: Summary data for the three PPP cohorts

|  | **UK** | **HUNT** | **FinnGen** |
| --- | --- | --- | --- |
| *Clinical Presentation* |  |  |  |
| Age of onset: Median (IQR) | 46 (35-55) | 56 (50-65) | 53 (42-60) |
| Concurrent plaque psoriasis: n (%) | 82 (28.5) | 15 (6.7) | 196 (19.5) |
| Psoriatic Arthritis: n (%) | 29 (10.1) | 56 (24.9) | 121 (12.0) |
| *Sex* |  |  |  |
| Female: n (%) | 228 (79.2) | 182 (80.9) | 718 (71.4) |
| Male: n (%) | 60 (20.8) | 43 (19.1) | 288 (28.6) |
| *Smoking status* |  |  |  |
| Current/former smoker: n (%) | 189 (65.7) | 151 (67.1) | NA |
| Never-smoker: n (%) | 20 (6.9) | 7 (3.1) | NA |
| Unknown: n (%) | 79 (27.4) | 67 (29.8) | 1006 (100) |

IQR, interquartile range; NA, not available

**Table E2**: Results obtained in individual cohorts for genome-wide significant and suggestive association signals.

|  |  |  | **UK** | | **HUNT** | | **FINNGEN** | | **Meta-analysis** | |
| --- | --- | --- | --- | --- | --- | --- | --- | --- | --- | --- |
| **rsID** | **Position** | **EAF/NEAF** | **P value** | **OR** | **P value** | **OR** | **P value** | **OR** | **P value** | **OR** |
| rs61802325 | 1:161,588,097 | A/G | 1.00 x 10^-01^ | 1.24 | 1.31 x 10^-01^ | 1.20 | 9.37 x 10^-08^ | 1.29 | 1.60 x 10^-08^ | 1.28 |
| rs887467 | 6:31,141,664 | C/G | 5.99 x 10^-05^ | 0.70 | 6.18 x 10^-01^ | 0.92 | 5.24 x 10^-09^ | 0.77 | 2.87 x 10^-11^ | 0.75 |
| rs73236841 | 4:37,911,079 | A/C | 1.11 x 10^-01^ | 0.80 | 1.30 x 10^-03^ | 0.59 | 1.25 x 10^-03^ | 0.84 | 4.79 x 10^-06^ | 0.78 |
| rs3798130 | 5:132,042,146 | T/C | 2.21 x 10^-01^ | 1.17 | 3.67 x 10^-02^ | 1.30 | 8.47 x 10^-06^ | 1.23 | 6.41 x 10^-07^ | 1.27 |
| rs4075959 | 5:176,784,612 | A/G | 3.64 x 10^-01^ | 1.10 | 3.04 x 10^-03^ | 1.36 | 4.83 x 10^-05^ | 1.21 | 1.10 x 10^-06^ | 1.26 |
| rs9487605 | 6:111,582,885 | A/G | 3.51 x 10^-02^ | 1.23 | 3.93 x 10^-01^ | 1.10 | 3.52 x 10^-05^ | 1.21 | 4.15 x 10^-06^ | 1.23 |
| rs2097442 | 6:32,422,191 | A/G | 2.29 x 10^-03^ | 1.22 | 1.30 x 10^-01^ | 1.24 | 3.44 x 10^-05^ | 1.22 | 1.43 x 10^-07^ | 1.29 |
| rs10950151 | 7:68,306,574 | T/C | 5.44 x 10^-01^ | 1.17 | 3.63 x 10^-05^ | 3.66 | 9.52 x 10^-04^ | 1.33 | 4.54 x 10^-06^ | 1.54 |
| rs1990107 | 7:84,724,076 | T/C | 2.58 x 10^-01^ | 1.39 | 9.42 x 10^-03^ | 2.61 | 5.21 x 10^-06^ | 1.71 | 1.75 x 10^-07^ | 2.00 |
| rs11793564 | 9:111,534,315 | A/G | 1.42 x 10^-01^ | 1.18 | 1.49 x 10^-02^ | 1.30 | 1.47 x 10^-04^ | 1.21 | 2.87 x 10^-06^ | 1.25 |
| rs4246905 | 9:117,553,249 | T/C | 4.98 x 10^-02^ | 0.81 | 2.29 x 10^-02^ | 0.78 | 6.47 x 10^-05^ | 0.79 | 6.16 x 10^-07^ | 0.78 |
| rs9666271 | 11:85,879,769 | T/C | 8.76 x 10^-02^ | 1.81 | 4.51 x 10^-01^ | 1.20 | 1.27 x 10^-05^ | 1.93 | 4.58 x 10^-06^ | 1.64 |
| rs860876 | 12:25,158,319 | T/G | 2.42 x 10^-03^ | 1.41 | 2.91 x 10^-02^ | 1.30 | 2.49 x 10^-03^ | 1.19 | 4.14 x 10^-06^ | 1.30 |
| rs112872175 | 17:80,589,968 | G/GATAA | 5.68 x 10^-03^ | 1.43 | 1.89 x 10^-02^ | 1.37 | 6.31 x 10^-04^ | 1.21 | 1.06 x 10^-06^ | 1.31 |
| rs4817988 | 21:40,468,838 | A/G | 9.21 x 10^-02^ | 0.83 | 5.08 x 10^-04^ | 0.69 | 1.13 x 10^-03^ | 0.84 | 2.15 x 10^-06^ | 0.79 |

EAF, effect allele frequency; NEAF, non-effect allele frequency; OR, odds ratio. ^1^Coordinates refer to GRCh37.

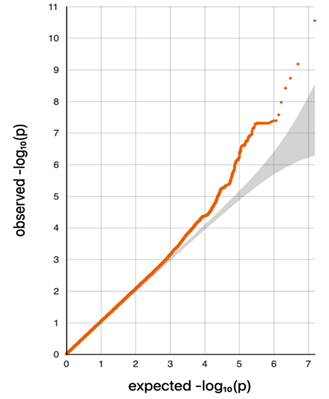

**Figure E1**: Meta-analysis quantile-quantile plot showing that the distribution of P-values did not significantly deviate from that expected under the null hypothesis (shaded in grey).

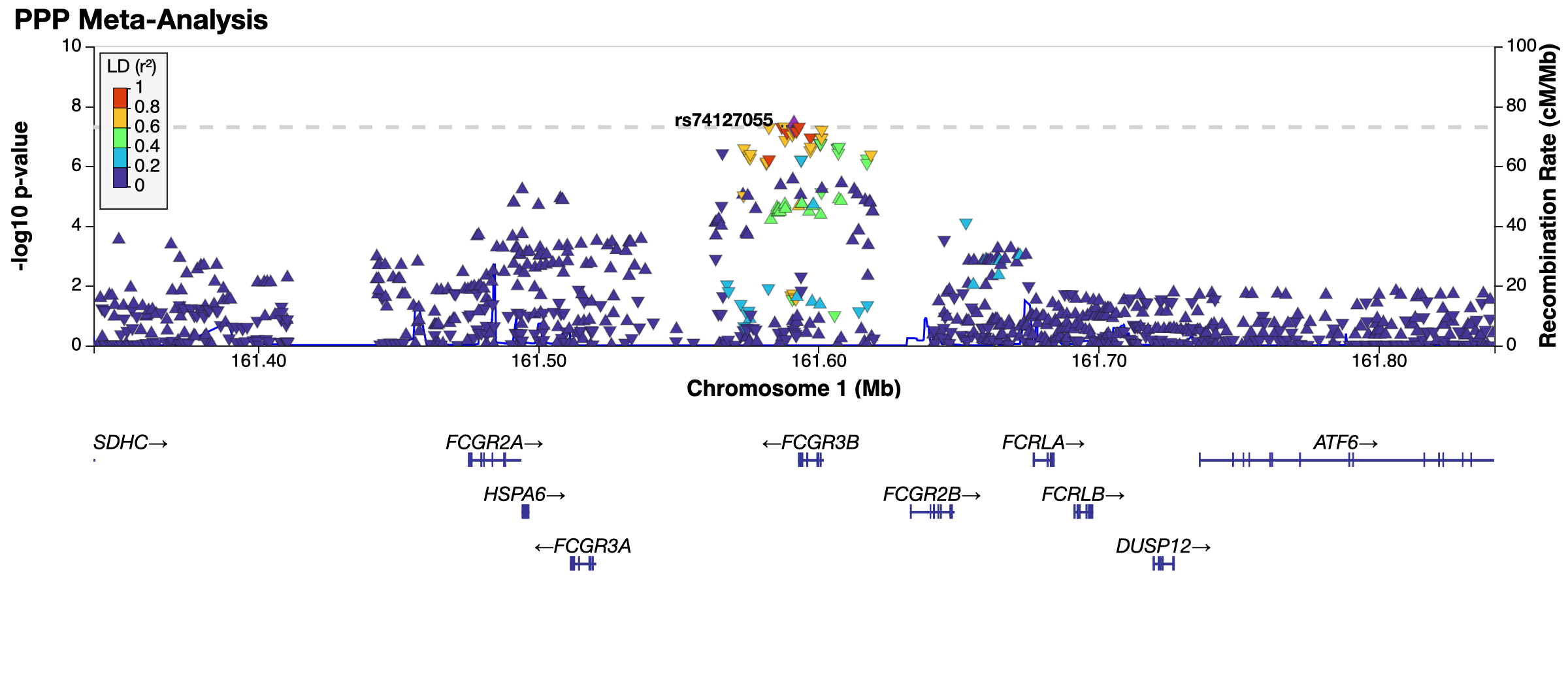

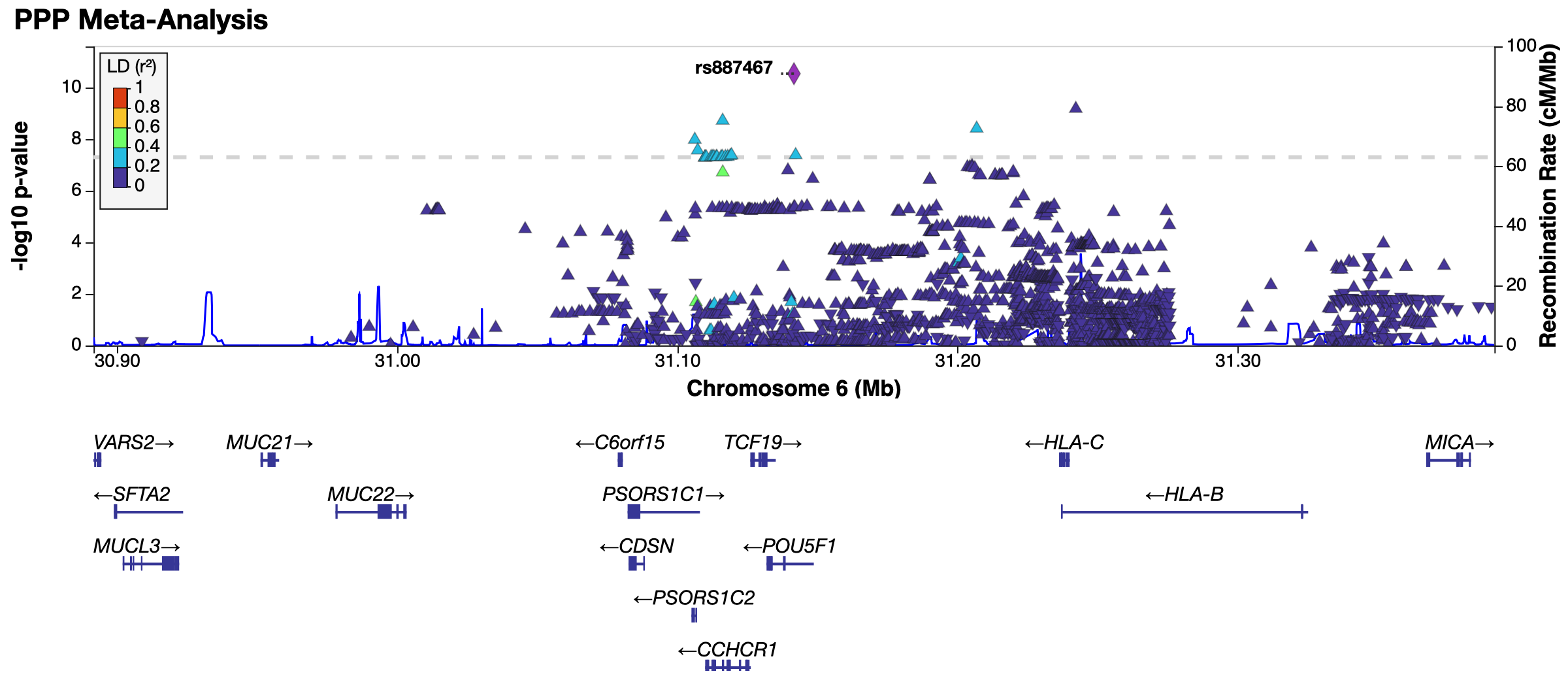
**Figure E2**: Regional association plots for the 1q23 and 6p21 susceptibility loci. P-values and recombination rates are plotted against chromosomal coordinates and gene positions.

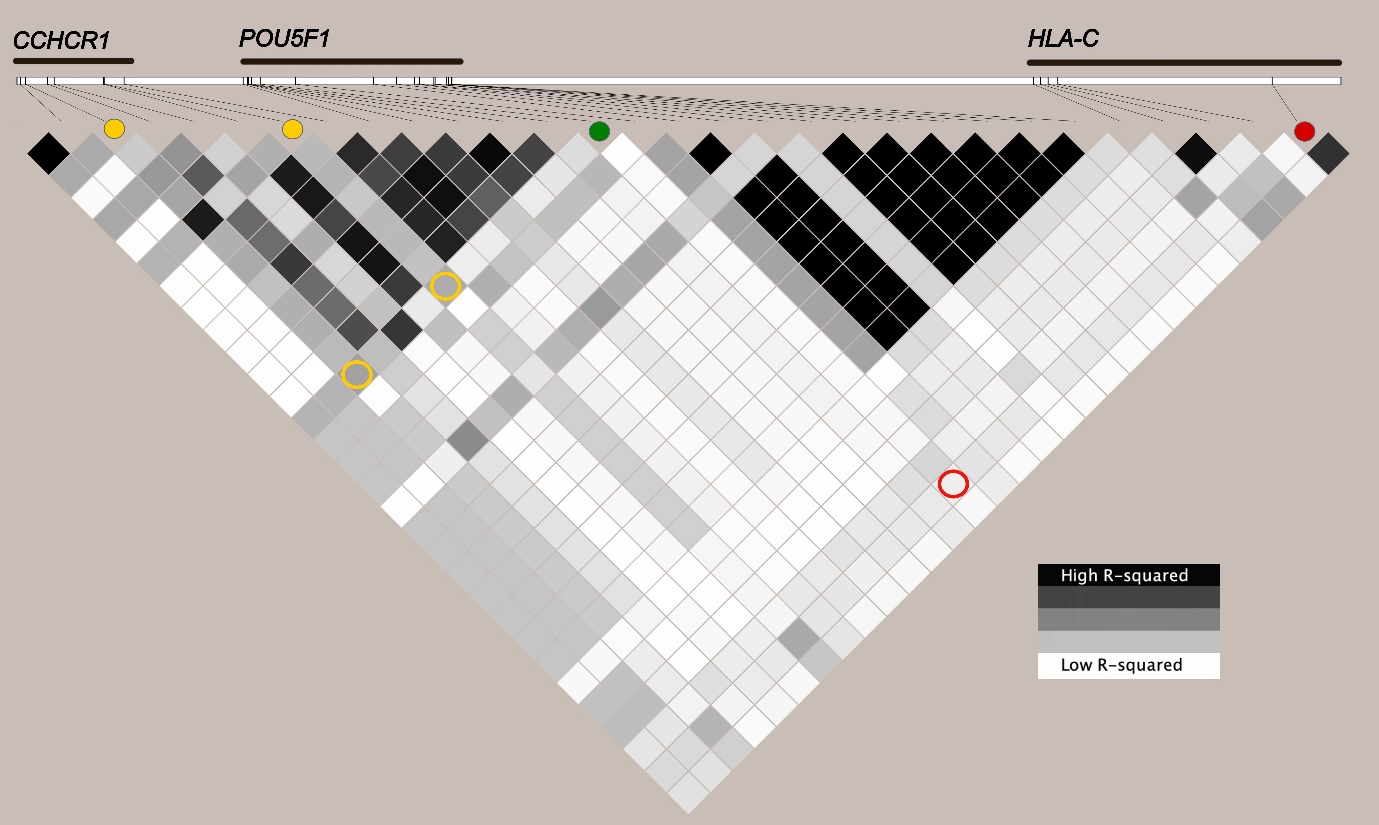

GBR

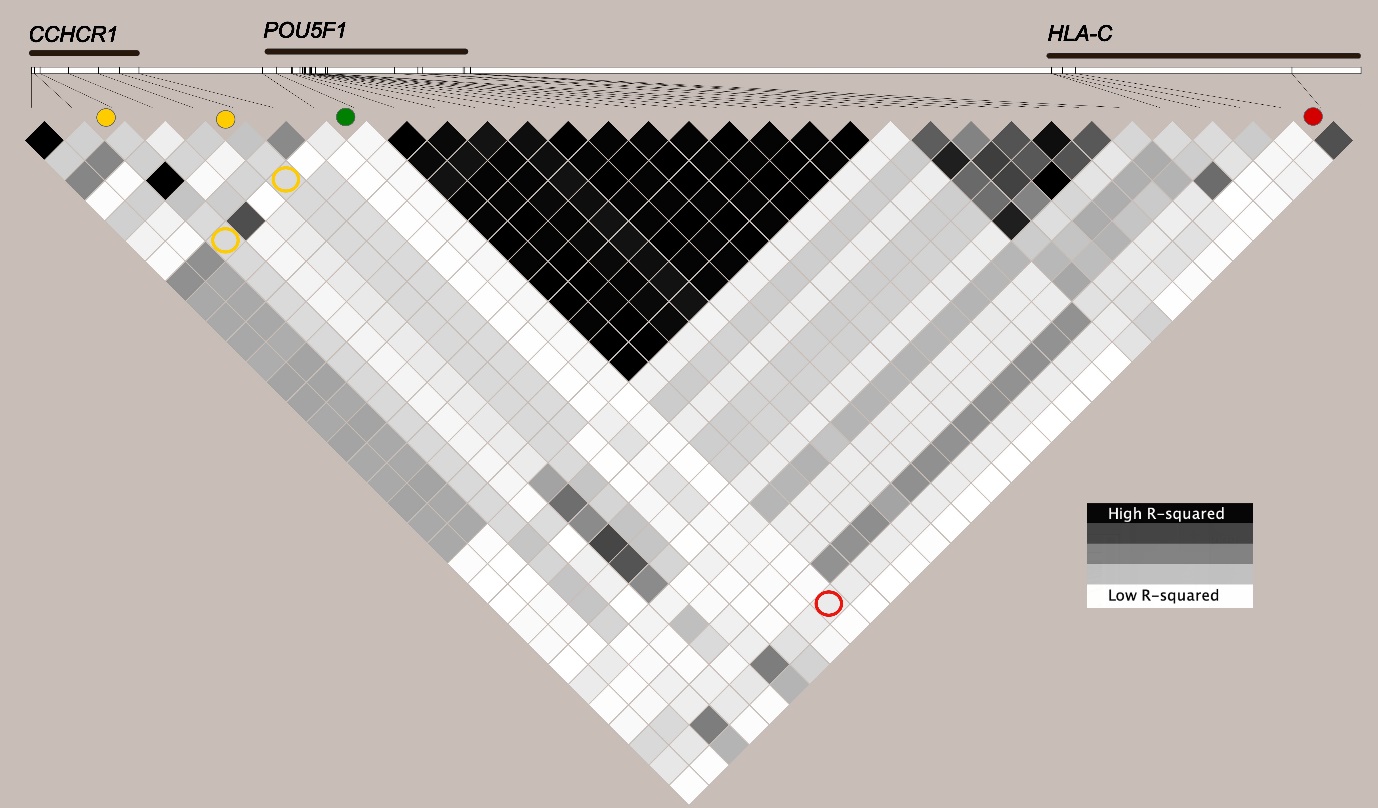

FIN

**Figure E3:** LD conservation plots showing pairwise r^2^ between SNPs genotyped by the 1,000 Genomes Project in British (GBR) and Finnish (FIN) individuals. The white bar at the top of each plot illustrates the position of the examined SNP with respect to *CCHCR1, POU5F1* and *HLA-C*. The coloured circles show that the lead SNP from the 6p21 region (green dot) is not in LD with the *HLA-Cw*0602* proxy SNP (red dot) or the *CCHCR1* SNPs previously associated with psoriasis (yellow dots).

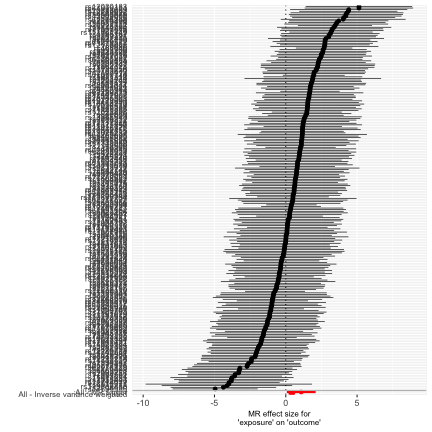

**Figure E4**: MR sensitivity analysis. Forest plot of the 240 smoking initiation SNPs associated with risk of PPP. The x-axis shows the MR effect size.
